# Pre-event psychiatric states predict trajectories of stress symptoms in Japan

**DOI:** 10.1101/2024.01.14.23300571

**Authors:** Fumiya Nakai, Taiki Oka, Nao Kobayashi, Masaaki Tanichi, Misa Murakami, Toshitaka Hamamura, Masaru Honjo, Yuka Miyake, Kentaro Ide, Aurelio Cortese, Masanori Nagamine, Takatomi Kubo, Toshinori Chiba

## Abstract

Recent time-dependent analyses of stress-related disorders have identified heterogeneity of trajectories and their modifying factors. While psychiatric patients are vulnerable to stress events, it is unclear how psychiatric conditions modulate subsequent stress symptoms. Using a longitudinal online survey from pre- to post-COVID-19 pandemic (n = 3815) and a latent growth mixture model, we identified four trajectories of stress symptoms: resilient, chronic, mild chronic, and early response. Though depression/anxiety was identified as a specific risk factor for the early-response trajectory, general psychiatric burden and social withdrawal were identified as common risk/protective factors. Further, we estimated pre-pandemic latent stress symptoms to determine the predictability of the stress symptoms. The chronic group showed significantly higher latent stress symptoms scores than the mild-chronic/early-response groups, both of which were significantly higher than the resilient group. These results suggest that prior psychiatric conditions may be considered for the prevention and treatment of maladaptive stress responses.

## INTRODUCTION

The COVID-19 pandemic has been a global calamity, and it offered a unique test case to investigate psychiatric resilience and the effects of stress exposure (Barzilay et al., 2020). Exposure to a stressful/traumatic event can cause severe mental problems, including severe stress symptoms, as have been observed in the context of the COVID-19 pandemic (Barzilay et al., 2020; Brülhart et al., 2021; Pierce et al., 2021). Recently, we found the strong temporal specificity of the association between stress symptoms toward the event and the suicide rate (Chiba et al., 2023). The finding supports that stress symptoms could thus be considered reliable surrogate endpoints for increased suicide that are highly heterogeneous across countries, cultures, and demographics (Brown et al., 1986; Harris et al., 2010; Judkins et al., 2020; Lantos & Nyári, 2024; Nagamine et al., 2020; Pirkis et al., 2021). Prevention of suicide can be progressed by understanding how stress reactions develop.

Recent research has revealed trajectories of the stress symptoms after the event exposures, especially PTSD after traumatic events (Andersen et al., 2014; Bonanno et al., 2012; Bonanno & Mancini, 2012; Dickstein et al., 2010; Karstoft et al., 2015; Orcutt et al., 2004; Saito et al., 2022). Trajectories of such stress symptoms are usually classified into four types: resilient, chronic, recovery, and delayed-onset (Andersen et al., 2014; Bonanno et al., 2012; Dickstein et al., 2010; Karstoft et al., 2015). The resilient type shows stable low or no PTSD severity. The chronic type shows consistently high severity. The recovery type shows rapid exacerbation that subsequently improves. The delayed-onset type shows low-level symptoms in the initial phase, which become exacerbated several months after the stressful trauma exposure (Andrews et al., 2007).

Trajectories of stress symptoms are shaped by a complex interplay of risk and protective factors, including personality, demographics, and coping strategies (Bonanno & Mancini, 2012; Dohrenwend et al., 2006). Among these, prior psychiatric conditions hold particular significance (Bomyea et al., 2012; Bonanno et al., 2023; Pan et al., 2021), but are poorly understood due to their bidirectional relationship with stress. Stress exacerbates these conditions, which, in turn, intensify stress symptoms, creating a reinforcing loop. This underscores the need for pre-event psychiatric data, which is rarely available since most patients seek care post-trauma. Previous research has addressed this limitation using data collected years before the stressful event (Solomon et al., 2021), yet the impact of near-term pre-event data remains underexplored. Moreover, the existing literature indicated the limited capacity to predict the stress symptom trajectories because they failed to capture heterogeneous aspects, only relying on a few pre-existing psychopathologic measures (Bonanno et al., 2023). Here, we bridge this gap by analyzing longitudinal mental health data collected just before and after the onset of the COVID-19 pandemic.

### Understanding latent stress symptoms prior to specific events

Stress symptoms often arise within a complex interplay of psychiatric conditions, which frequently share genetic and neurobiological roots. For instance, PTSD’s strong association with depression and anxiety underscores these connections. Cognitive biases, extensively documented in psychological research (Harding et al., 2004; Kremer et al., 2020; Noworyta-Sokolowska et al., 2019), often distort perceptions, leading individuals to attribute enduring psychological burdens to recent stressors. These pre-existing burdens can be conceptualized as "latent stress symptoms," remaining unexpressed until exacerbated by life events. This study utilizes concurrent psychiatric assessments to model these latent stress symptoms, elucidating their critical role in shaping subsequent stress responses. By distinguishing pre-existing latent stress symptoms from event-driven exacerbations, this framework advances our understanding of stress symptomatology and enhances approaches to stress-related disorders. In this study, we examined the effects of psychiatric states and demographics sampled before the COVID-19 pandemic on trajectories of stress symptoms during the pandemic.

Further, using latent stress symptoms, we provide an intuitive interpretation of prognosis in stress symptoms from the pre-event data.

## TRANSPARENCY AND OPENNESS

### Preregistration

This study was not preregistered.

### Data, materials and online resources

The main summary statistical and materials data supporting this study’s findings are available in the manuscript and Supplementary Materials. Owing to company cohort data sharing restrictions, individual-level data cannot be publicly posted. However, data are available from the authors upon request and with permission of KDDI Corporation.

### Reporting

We report how we determined our sample size, all data exclusions, all manipulations, and all measures relevant to this study.

## METHODS

### Procedures and outcomes

The online survey was conducted by Macromill Inc.(Japan) as a large, longitudinal survey to investigate the mental health of the general Japanese adult population (see our previous studies (Chiba et al., 2023; Oka, Hamamura, et al., 2021; Oka, Kubo, et al., 2021) for details). The original panel survey was conducted in December 2019 (T0) before the identification of the first COVID-19 case in Japan (January 2020). In response to the outbreak of COVID-19, follow-up surveys of T0 participants were conducted in August 2020 (T1), December 2020 (T2), April 2021 (T3), August 2021 (T4), and December 2021 (T5). Invitations for the original survey were sent to 5955 individuals, of which 478 were excluded due to inconsistencies or contradictions in their answers. An additional 481 individuals were also excluded because of unreliable answers, such as using only the maximum or minimum rating in questionnaires, including reverse items. The sample size was determined in accordance with the original large-scale study, which aimed to examine psychopathological issues in a cohort of over 5,000 individuals from the general population during the COVID-19 pandemic. The original and follow-up research designs were approved by the Ethics Committees of the Advanced Telecommunications Research Institute International (ATR) (approval No. 21-195 for the original study & 21-749 for the follow-up study). This study was conducted in accordance with the guidelines of the Declaration of Helsinki. All participants read a full explanation of the study and gave informed consent before each survey.

Questionnaires were constructed from questions for psychiatric, demographic, and COVID-19 related items. Our survey included ten types of validated questionnaires for psychiatric disorders; PTSD (IES-R (Weiss, 2004)), major depressive disorder (CES-D (Radloff, 1977)), obsessive-compulsive disorder (OCI (Foa et al., 1998)), internet-related problems (CIUS (Meerkerk et al., 2009)), attention-deficit/hyperactivity disorder (ADHD) (ASRS (Kessler et al., 2005)), autistic spectrum disorder (AQ (Baron-Cohen et al., 2001)), social anxiety (LSAS-fear/avoid (Baker et al., 2002)), general anxiety (STAI-Y-state (Spielberger, 1983)), and alcohol-related problems (AUDIT (Saunders et al., 1993)). To assess stress symptoms severity, we used the IES-R, which is commonly used for PTSD assessment, to examine stress symptoms to COVID-19 (Husky et al., 2021). Although limitations have been noted in measuring stress symptoms with PTSD scales that are inherently event-related,(Muysewinkel et al., 2024) traumatic and stressful experiences that do not meet the A criteria are treated as mild/small trauma, which is known to cause symptoms similar to PTSD (Marx et al., 2024). We dealt with trajectories of stress symptoms to pandemics, which are positioned as mild trauma (Bridgland et al., 2021). Moreover, this approach facilitates comparison with previous research on trajectories, as the IES-R is frequently used in trajectory studies (Galatzer-Levy et al., 2018).

Collected demographic data included sex (women and men), age, job status (self-employed, employed, unemployed, and other), marital state, and household income per year (lowest; less than four million yen, 2nd; four∼six million yen, 3rd; six∼eight million yen, 4th; eight∼ten million yen, Highest; more than ten million yen, and Missing) and they were analyzed as pre-COVID demographics.

### Trajectory analysis using the Latent Growth Mixture Model

In this study, we assumed that the global heterogeneity of stress symptom trajectories can be explained by a set of homogeneous trajectories. Consistent with previous studies, we used a latent growth mixture model (LGMM) (Muthén, 2004; Muthén & Muthén, 2000) to identify latent class trajectories of stress symptoms measured by IES-R from T1 to T5 during the COVID-19 pandemic. The “*lcmm*” package^45^ in R version. 4.1.0 was used to identify the latent classes. The classification was performed under all parameter conditions (6×3: the number of latent classes (1 to 6 class) × the model function (linear, quadratic and exponential)). The optimal condition was determined according to the following measures. Commensurate with recommendations in a previous study (Nylund et al., 2007), we relied on three types of measures to determine the best model for clustering: AIC (Akaike information criterion), BIC (Bayesian information criterion), and sample-size-adjusted BIC. A grid search approach with 100 iterations was used to estimate optimal values of model parameters, and 100 repetitions were applied to achieve stable results. After repetition, the optimal parameter was determined using the maximum likelihood method. After estimating optimal model parameters, the membership probability was calculated for each participant to assign a class label. In the model with the quadratic function, which showed the lowest BIC for all class numbers, the Vuong-Lo-Mendell-Rubin Likelihood Ratio Test (VLMR) was used to compare the model fitting between models with adjacent class numbers.

### Multinomial logistic regression analysis

We used a multinomial logistic regression model to identify risk/protective factors for each trajectory class. Prior to applying the model, the nine psychiatric scores of all participants were compressed into four-dimensional data using principal component analysis (PCA) on T0 data (Oka, Kubo, et al., 2021; Wold et al., 1987). Then, according to the estimated loading of the top four PCs, orthogonal transformation was applied, and the scores of each participant at each time point were converted into four-dimensional orthogonal scores. Then, logistic regression was performed using class assignment as a dependent variable and compressed psychiatric scores at T0 and other demographics (sex, age, income, employment status, and marital status) as independent variables. The relative risk b from the resilient group for each independent variable was estimated for each trajectory group. A *p*-value less than 0.05 indicates statistical significance, and *p*-values were adjusted by Bonferroni correction. We fitted the model using the *Statistical Machine Learning Toolbox* in *Matlab*.

### Estimation of latent stress symptoms score

We estimated the latent stress symptoms score by using multiple psychiatric scores collected prior to COVID-19 in multiple ridge regression analysis (Bishop, 2006; Vapnik et al., 1996). First, all data from T1 to T5 were randomly split into training and test sets in a 9:1 ratio. To obtain reliable results excluding sampling bias, we repeated this random splitting procedure 10 times, thus creating ten randomly sampled datasets. For each dataset, both training and test data were normalized based on the mean and variance of the training dataset. Second, Ridge regression was applied to analyze the linear association between stress symptom scores and nine other psychiatric scores from T1 to T5 in each dataset. Training data were further divided into training and validation data with a 5-fold cross-validation approach to tune the regularization parameter in each randomly sampled dataset. The optimal penalty coefficient was searched within the range of 0.001 to 10,000 using logarithmic increments of 10. In each cross-validation, regression and penalty coefficients were determined based on the minimization of the cost function estimated with the validation dataset. Successively, optimal parameters were selected based on the minimization of the cost function among the folds of cross-validation. The model was evaluated using the test data based on the adjusted-*R*^2^ metrics in each dataset, and we reported the mean score of the adjusted-*R*^2^ metrics. Using the final model estimated, the latent stress symptoms of test data were calculated from the nine psychiatric scores at T0 in each dataset. Finally, we concatenated the test data from each dataset with the estimated latent stress symptoms and used the values for the subsequent statistical analysis. The difference in the mean latent stress symptoms score for each trajectory group was tested by one-way ANOVA, with post-hoc Tukey tests.

Furthermore, we evaluated the model’s applicability across time points with a similar approach. Specifically, we performed leave-one-time-point-out cross-validation. In this validation, the model was trained on scores at four time points, except fixed time point Tx (selected between T1 and T5), and the scores at Tx were used as test data.

## RESULTS

### Descriptive results

The number of participants who contributed to the data at T0 was 3815 (51.3% male and 48.7% female), and the mean age was 46.3 (45.9-46.6 for 95% CI) years (see Table 1). The statistical test revealed a significant decrease in stress symptoms severity from T1 to T2-T5, from T2 to T5, and from T3 to T5 (one-way ANOVA: *p* <.001, F_4,12594_ = 60.69, η^2^ = 0.02, post-hoc Tukey test: all *p*<.001).

**Table 1.** Demographics of the survey.

| Characteristic | Time Point of Survey |  |  |  |  |  |
| --- | --- | --- | --- | --- | --- | --- |
|  | T0 | T1 | T2 | T3 | T4 | T5 |
| Participants, No. | 3815 | 3508 | 2680 | 2562 | 2202 | 1806 |
| Age (95%CI) | 46.3 (45.9-46.6) | 46.4 (46.1-46.8) | 47.0 (46.6-47.5) | 47.1 (46.7-47.6) | 47.7 (47.3-48.2) | 47.8 (47.3-48.3) |
| Gender, % |  |  |  |  |  |  |
| Male | 51.3 | 51.7 | 52.6 | 52.8 | 54.8 | 55.5 |
| Female | 48.7 | 48.3 | 47.4 | 47.2 | 45.2 | 44.5 |
| Income (million), % |  |  |  |  |  |  |
| -2 | 5.9 | 6.0 | 5.9 | 5.8 | 5.2 | 5.3 |
| 2-4 | 18.3 | 18.4 | 18.3 | 18.8 | 18.7 | 18.9 |
| 4-6 | 21.6 | 21.6 | 21.8 | 21.4 | 21.8 | 21.9 |
| 6-8 | 17.7 | 17.5 | 18.2 | 18.5 | 18.7 | 18.0 |
| 8-10 | 8.9 | 9.0 | 9.7 | 9.4 | 10.5 | 11.0 |
| 10-12 | 3.9 | 3.9 | 4.0 | 4.4 | 4.6 | 4.5 |
| 12-15 | 2.3 | 2.3 | 2.5 | 2.5 | 2.6 | 2.6 |
| 15-20 | 1.4 | 1.4 | 1.4 | 1.4 | 1.6 | 1.7 |
| 20- | 0.8 | 0.8 | 0.7 | 0.8 | 0.9 | 0.8 |
| N/A | 19.1 | 19.0 | 17.5 | 17.0 | 15.4 | 15.4 |
| Employment, % |  |  |  |  |  |  |
| employed | 77.7 | 77.8 | 77.7 | 77.9 | 78.8 | 78.5 |
| unemployed | 22.3 | 22.2 | 22.3 | 22.2 | 21.2 | 21.5 |
| Married, % |  |  |  |  |  |  |
| married | 64.6 | 64.8 | 65.2 | 64.3 | 65.1 | 65.1 |
| unmarried | 35.4 | 35.2 | 34.8 | 35.7 | 34.9 | 34.9 |

### Latent growth mixture model analysis

A latent growth mixture analysis (LGMM) was used to identify the latent class trajectories of stress symptoms. For the LGMM analysis, all pairs of model functions and numbers of latent classes were tested and evaluated by statistical indicators. As a result, all three indicators (AIC, BIC, and adjusted-BIC) showed that a quadratic function with a four-class model offered the best fit (see Table S1). We excluded a five-class model with the quadratic function from the analysis because the number of classes converging in model fitting was less than the specified number. In other words, while we attempted to estimate a five-class model, one or more of the classes failed to converge, resulting in an invalid solution. The Vuong-Lo-Mendell-Rubin Likelihood Ratio Test (VLMR) was also performed for the fitted models with quadratic function to compare the model fitting for each class number, and the 4-class model was significantly better than the 3- and 5-class models (Class 3; *p* < 0.001, df = 15, Class 5; *p* <0.001, df = 19). Consistent with previous studies (deRoon-Cassini et al., 2010; Murphy & Smith, 2018; Steenkamp et al., 2012), four types of trajectory were named: resilient, chronic, mild chronic, and early response (Figure 1a). The resilient group comprised 2712 participants [71.1%], the chronic group consisted of 120 participants [3.2%], the mild-chronic group included 623 participants [16.3%], and the early response group consisted of 360 participants [9.4%] (Figure 1b). In the resilient group, a significant decrease was observed at T2 from T1 (*p* <.001, *t* = 5.25, df = 4418) and T5 from T4 (*p* <.001, *t* = 4.02, df = 2805). No significant change was observed in the chronic group at any time pair. In the mild chronic group, a significant decrease was observed at T4 from T3 (*p* <.001, *t* = 4.51, df = 740) and at T5 from T4 (*p* <.001, *t* = 4.77, df = 602). Finally, in the early response group, a significant decrease was observed at T2 from T1 (*p* <.001, *t* = 28.09, df = 579) and at T3 from T2 (*p* <.001, *t* = 5.66, df = 433), but a significant increase was also observed at T5 from T4 (*p* <.001, *t* = 5.19, df = 306) (all trajectories are shown in Figure 1c). While most results were consistent with previous studies, the delayed-onset trajectory was not identified in our study. This may be explained by the difference in the observation period, interval, and the event type (Bonanno & Mancini, 2012).

**Figure 1.**
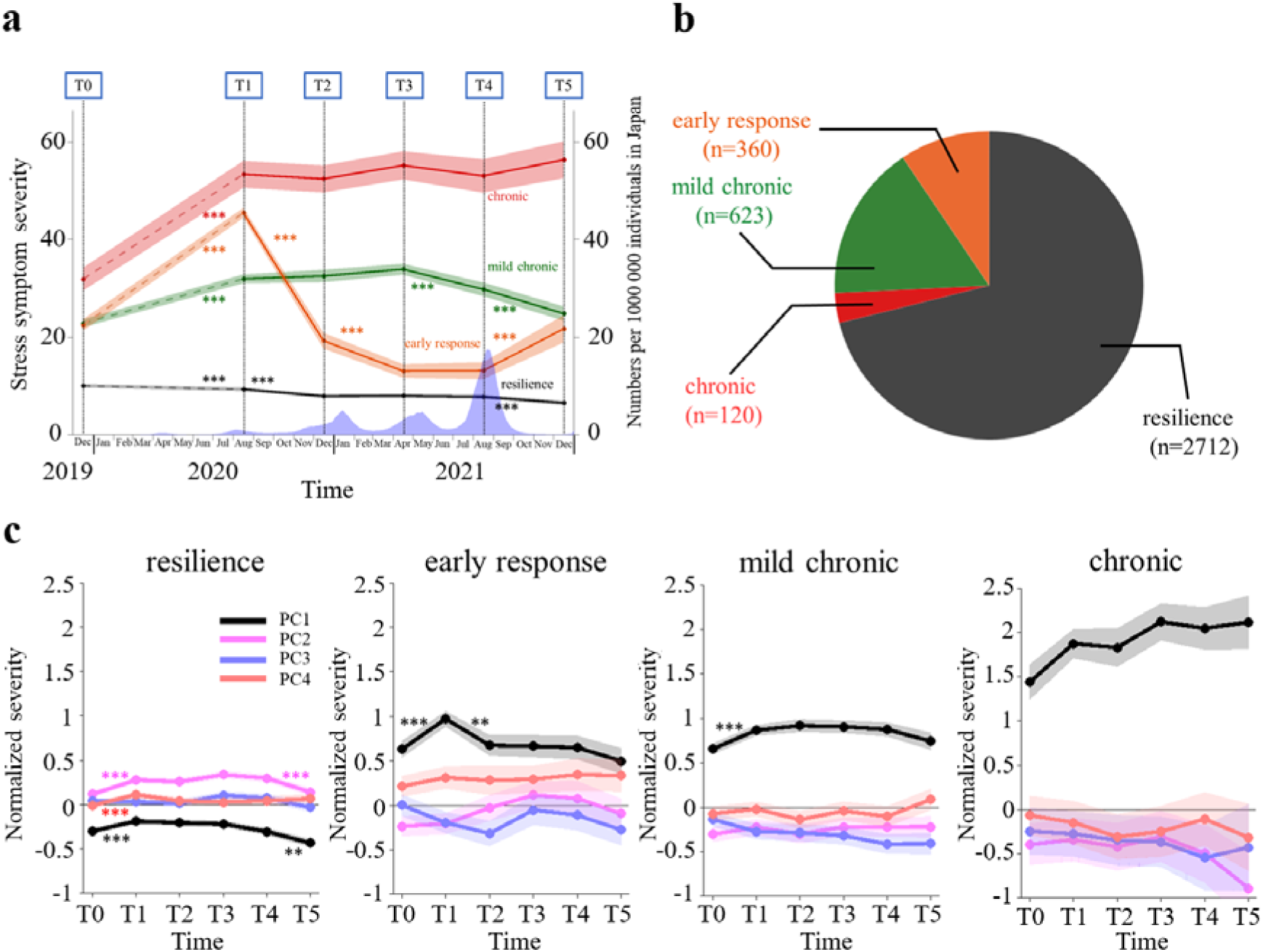
The trajectories of stress symptom scores and PC trajectories for each group **a.** Thick lines show mean stress symptom trajectories of the four groups estimated by LGMM, and thin lines show their 95% confidence intervals. Stress symptoms were normalized at T1, and T0 scores were estimated by Ridge regression. Each time transition was tested by unpaired t-test (* shows significant difference with Bonferroni correction (n=5)). The blue bars represent a moving average in back-and-forth seven days of new daily cases of COVID-19 per 100,000 Japanese residents. **b.** Population ratio of four estimated groups. **c.** Thick lines show mean trajectories of psychiatric items estimated by PCA for each of the four groups, and thin lines show their 95% confidence intervals. Each time transition was tested by ANOVA (* shows significant difference adjusted by Bonferroni correction (n=5)).

### Multinomial logistic regression analysis

Psychiatric scores at T0 were compressed by principal component analysis (PCA), and the top 4 components (25.3% for PC1, 13.9% for PC2, 11.2% for PC3, 10.9% for PC4, and 61.2% for all, Figure S1) were used in the sequential modifying factor analysis. Identified factors were consistent with the previous study^5^, and each PC was named in the same way: PC1 as a general psychiatric burden, PC2 as social withdrawal, PC3 as alcohol-related problems, and PC4 as depression/anxiety.

Risk factors for each group were identified by multinomial logistic regression (MLR) with the resilient group as a reference (multiple comparison was corrected by the Bonferroni method, Figure 2). As a result, a general psychiatric burden component (PC1) was identified as a significant risk factor for all groups(chronic: *b* = 2.16; 95% CI [1.89-2.42], *p* <.001, *t* = 15.87, df = 9234, mild chronic: *b* = 1.27; 95% CI [1.14-1.40], *p* <.001, *t* = 19.13, df = 9234, early response: *b* = 1.24; 95% CI [1.08-1.39], *p* <.001, *t* = 15.62, df = 9234). Also, a social withdrawal component (PC2) was identified as a significant protective factor for all groups (chronic: *b* = -0.70; 95% CI [-0.90-0.49], *p* <.001, *t* = -6.57, df = 9234, mild chronic: *b* = -0.51; 95% CI [-0.62-0.40], *p* <.001, *t* = -8.88, df = 9234, early response: *b* = -0.43; 95% CI [-0.56-0.30], *p* <.001, *t* = -6.39, df = 9234). An alcohol-related problem component (PC3) was identified as a significant protective factor for the chronic group (*b* = -0.30; 95% CI [-0.49- 0.10], *p* <.01, *t* = -3.02, df = 9234) and the mild chronic group (*b* = -0.23; 95% CI [-0.34-0.13], *p* <.001, *t* = -4.30, df = 9234). Depression/anxiety component (PC4) was also identified as a significant risk factor for the early response group (*b* = 0.23; 95% CI [0.10-0.35], *p* = .005, *t* = 3.55, df = 9234). While young age and unmarried status also showed a tendency as a risk factor for the chronic group, which was consistent with the previous result (Pierce et al., 2021), no significant difference was identified (see Table S2 for the details of statistical values.

**Figure 2.**
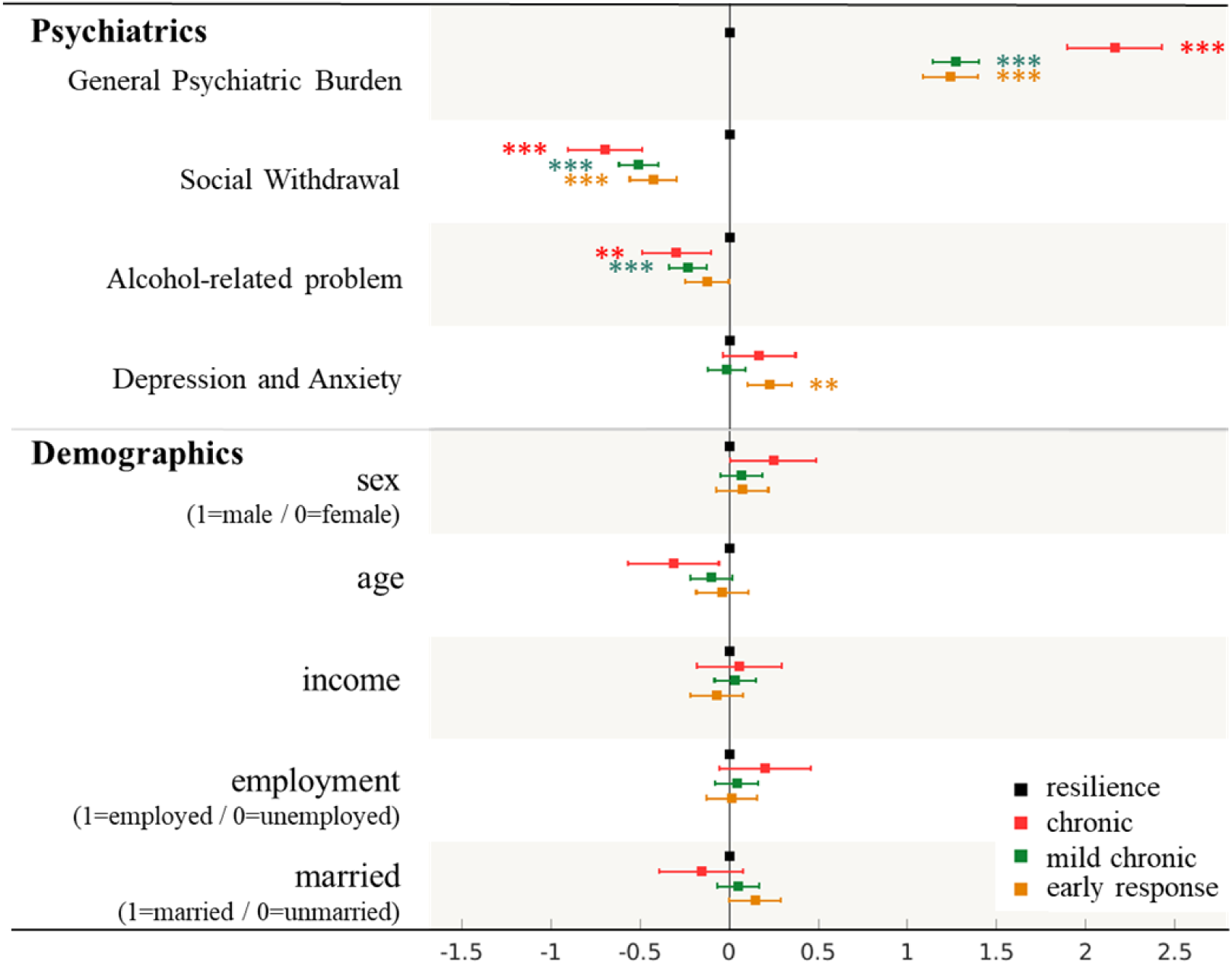
Multinomial logistic regression analysis showing risk/protective factors for each stress symptom group Multinomial logistic regression (MLR) analysis shows risk/protective factors for each stress symptom group. Psychiatric items were estimated by principal component analysis (PCA) based on scores of nine other psychiatric symptoms. Demographic items were selected as pre-existing demographics before the trigger event. Each dot shows coefficients of the MLR, and the line shows the 95%-ile. Scores of the resilient group were used as a reference, and * shows a significant difference from the reference (adjusted by Bonferroni correction (n=3)). *** *p_corr_*< 0.001, *p_corr_* < 0.01

### Latent stress symptoms estimation

The latent stress symptoms score of the chronic group (30.7; 95% CI [28.8-32.7]) was significantly higher than those of the mild chronic (22.8; 95% CI [22.1-23.6]), early-response (22.2; 95% CI [21.1-23.2]) and resilient groups (10.7; 95% CI [10.4-11.0]), and scores of mild chronic and early-response groups were significantly higher than that of the resilient group (one-way ANOVA, *p* <.001, *F_3,3806_* = 565.1, η*^2^* = 0.52; posthoc Tukey test chronic versus mild chronic, early-response, and resilient *p* <.001; resilient versus mild chronic, early response *p* <.001). No significant difference was found between the mean scores of the mild chronic and early-response groups.

We also tested the estimation validity of latent stress symptoms score by using leave-one-time-point-out cross-validation analysis. The mean adjusted-*R*^2^ score of the estimation model showed 0.525 at training and 0.436 at testing. The leave-one-time-point-out cross-validation analysis showed higher than 0.5 adjusted-*R*^2^ score for all Tx conditions at training and higher than 0.4 adjusted-*R*^2^ score for all Tx conditions at test (see details in Supplementary materials).

## DISCUSSION

In this study, we conducted an online longitudinal survey that included event-related stress symptoms over a COVID-19 pandemic period of 1.5 years since the event’s exposure. Our LGMM trajectory analysis identified four stress symptoms trajectory types: resilient, chronic, mild chronic, and early response. We then demonstrated how these prognoses are modulated by psychiatric conditions prior to the pandemic. For demonstration purposes, we further estimated the latent stress symptoms score before the pandemic, which we assume corresponds to the score that would have arisen during the pandemic if this psychiatric exacerbation had not occurred. We found that the higher latent stress symptoms were associated with higher stress symptoms during the pandemic, with more pronounced exacerbation in stress symptoms.

This analysis highlights that pre-pandemic psychiatric conditions influence stress trajectories both distinctly and broadly, guiding two intervention strategies: tailored, trajectory-specific care and general approaches. Depression and anxiety, key risk factors for the early response group, may reflect transient and normal stress responses. For these cases, symptomatic psychotherapy, psychoeducation, and short-term pharmacotherapy (e.g., anxiolytics) may prove more effective than SSRIs, focusing on alleviating immediate distress. Normalizing these responses through education may further foster understanding and recovery, promoting resilience during stressful events (Bersani & Delle Chiaie, 2021; Cohen & Mannarino, 2015)^.^

In contrast to a specific risk factor, the social withdrawal/general psychiatric burden component emerged as a shared risk and protective factor across stress trajectories. While most individuals faced mental health declines during the pandemic (Brunoni et al., 2021; van der Velden et al., 2021), those predisposed to social isolation and lower sociability may have found psychological relief in reduced social demands. This finding highlights the importance of personalizing public health strategies, such as social distancing policies, to respect individual inclinations toward social interaction. The general psychiatric burden component was also identified as a common risk factor.

This finding aligns with established observations indicating the vulnerability of psychiatric patients to stress.(Goh & Agius, 2010; Harris et al., 2010) From this perspective, those with high general psychiatric burden components may have experienced a more pronounced exacerbation of their psychological states. On the other hand, from the perspective that many psychiatric disorders arise from a single common factor (Caspi & Moffitt, 2018), stress-induced psychiatric symptoms may be attributable to emotional malaise that existed before the event. Individuals may have interpreted their malaise that existed before the pandemic rather than that was induced by the pandemic. In such a scenario, the individual’s holistic problem, rather than the effect of the trigger event, should be resolved to treat stress symptoms. To identify which perspective better explains the mechanism of stress symptom deterioration, it is necessary to separate psychiatric problems existing prior to the event—latent stress symptoms—from those exacerbated during the event.

To achieve this, we estimated the latent stress symptoms using pre-pandemic psychiatric conditions based on the correspondence between stress symptoms and psychiatric conditions during the pandemic. Here, we assume that latent stress symptoms correspond to symptoms attributed to psychiatric conditions prior to stressful events. Leave-one-time-out regression suggested that magnitudes in stress symptoms could be estimated from scores of other psychiatric disorders at other time points (mean adjusted-R^2^ = 0.411). The ANOVA conducted on the mean latent stress symptoms for trajectory types revealed three levels: high for the chronic group, low for the resilient group, and intermediate for the others. This finding suggests that individuals with higher latent stress symptoms are more likely to exhibit elevated stress symptoms following a stressful life event. During prolonged stressors, such as the COVID-19 pandemic, these heightened stress levels can establish a new baseline, creating a feedback loop where elevated symptoms perpetuate and intensify over time. This cyclical effect highlights the importance of early intervention to prevent the compounding of stress symptoms throughout extended periods of adversity. While the precise interpretation of the latent stress symptoms requires further discussion and clarification, our proposal represents an initial step toward addressing the intricate relationship between stress vulnerability and exacerbations triggered by the event.

There are several limitations in our study. First, though small classes could present challenges for replication (Nylund-Gibson & Choi, 2018), our chronic stress class captured only 3.2%. While we acknowledge the concern about the small size, the chronic stress group aligns with prior literature and clinical observations (Galatzer-Levy et al., 2018), where a small subset of individuals demonstrate persistent and severe symptoms over time. This group’s inclusion provides critical insights into heterogeneity within the sample. Second, it is also important to verify whether the results of our study can be generalized to other types of stressful experiences; therefore, it is necessary to apply the same analysis to other cases. Third, our measurements were taken every three months, which means we did not capture specific stress events occurring over shorter spans, which may have led to inconsistent results compared to the existing literature, e.g., a lack of a delayed onset group. While the results have a variety of implications for stress research, they should be interpreted with caution in comparison to previous studies at various time and population scales (Schäfer et al., 2022).

In conclusion, we identified four types of stress symptoms with different modifying factors. We also found that these four trajectories were partly predictable, even before the onset of the stressful event. These findings may help develop personalized care according to the psychiatric states underlying their pathogenesis.

## Data Availability

The main summary statistical data that support the findings of this study are available in the Supplementary Data. Owing to company cohort data sharing restrictions, individual-level data cannot be publicly posted. However, data are available from the authors upon request and with permission of KDDI Corporation.

## Corresponding Author

Toshinori Chiba, Ph.D., MD, Department of Decoded Neurofeedback, Computational Neuroscience Laboratories, Advanced Telecommunications Research Institute International, 2-2-2 Hikaridai, Seika-cho, Soraku-gun, Kyoto, 619-0288, JAPAN, and Takatomi Kubo, Ph.D., MD, Department of Science and Technology, Nara Institute of Science and Technology, 8916-5 Takayama-cho, Ikoma, Nara, 630-0192, JAPAN.

## Author Affiliations

Department of Decoded Neurofeedback, Computational Neuroscience Laboratories, Advanced Telecommunications Research Institute International (ATR), Kyoto (FN, TK, TO, MM, KI, CA, TC), Department of Science and Technology, Nara Institute of Science and Technology (NAIST), Kyoto (FN, TK), Department of Neuropsychiatry, Faculty of Life Sciences, Kumamoto University, Kumamoto (TO), Clinical Psychology, Graduate School of Human Sciences, Osaka University, Suita (TO), Life Science Laboratories, KDDI Research, Inc., Tokyo (NK, TH, MH, YM), The Department of Psychiatry, Self-Defense Forces Hanshin Hospital, Kawanishi (MT, TC), Division of Behavioral Science, National Defense Medical College Research Institute, Tokorozawa (MN).

## Author Contribution

FN, TO, TK, and TC had full access to all of the study’s data and are responsible for the integrity of the work.

*Conceptualization*: FN, TO, TK, TC. *Investigation*: TO, TH, NK, MM, MH, YM, TC. *Methodology*: FN, TO, MT, TH, MN, KI, TK, TC.

*Validation*: TO, TK, TC

*Formal analyses*: FN, TO, TK, TC.

*Writing – Original Draft Preparation*: FN, TO, AC, TK, TC.

*Writing – Review & Editing*: All authors.

*Project Administration*: TO, TH, NK, TK, TC

## Acknowledgements

We thank Rumi Yorizawa (The Department of Decoded Neurofeedback, Computational Neuroscience Laboratories, Advanced Telecommunications Research Institute

International, Kyoto), and Miho Nagata (The Department of Decoded Neurofeedback, Computational Neuroscience Laboratories, Advanced Telecommunications Research Institute International, Kyoto) for data collection and organization.

## References

1. Andersen, S. B., Karstoft, K.-I., Bertelsen, M., & Madsen, T. (2014). Latent trajectories of trauma symptoms and resilience: the 3-year longitudinal prospective USPER study of Danish veterans deployed in Afghanistan. The Journal of Clinical Psychiatry, 75(9), 1001–1008.

2. Andrews, B., Brewin, C. R., Philpott, R., & Stewart, L. (2007). Delayed-onset posttraumatic stress disorder: a systematic review of the evidence. The American Journal of Psychiatry, 164(9), 1319–1326.

3. Baker, S. L., Heinrichs, N., Kim, H.-J., & Hofmann, S. G. (2002). The liebowitz social anxiety scale as a self-report instrument: a preliminary psychometric analysis. Behaviour Research and Therapy, 40(6), 701–715.

4. Baron-Cohen, S., Wheelwright, S., Skinner, R., Martin, J., & Clubley, E. (2001). The autism-spectrum quotient (AQ): evidence from Asperger syndrome/high-functioning autism, males and females, scientists and mathematicians. Journal of Autism and Developmental Disorders, 31(1), 5–17.

5. Barzilay, R., Moore, T. M., Greenberg, D. M., DiDomenico, G. E., Brown, L. A., White, L. K., Gur, R. C., & Gur, R. E. (2020). Resilience, COVID-19-related stress, anxiety and depression during the pandemic in a large population enriched for healthcare providers. Translational Psychiatry, 10(1), 291.

6. Bersani, F. S., & Delle Chiaie, R. (2021). The end method: Normalization. In M. Biondi (Ed.), Empathy, Normalization and De-escalation (Vol. 147, pp. 57–64). Springer International Publishing.

7. Bishop, C. M. (2006). Pattern Recognition and Machine Learning (1st ed.). Springer.

8. Bomyea, J., Risbrough, V., & Lang, A. J. (2012). A consideration of select pre-trauma factors as key vulnerabilities in PTSD. Clinical Psychology Review, 32(7), 630–641.

9. Bonanno, G. A., Chen, S., & Galatzer-Levy, I. R. (2023). Resilience to potential trauma and adversity through regulatory flexibility. Nature Reviews Psychology, 2(11), 663–675.

10. Bonanno, G. A., & Mancini, A. D. (2012). Beyond resilience and PTSD: Mapping the heterogeneity of responses to potential trauma. *Psychological Trauma: Theory, Research*, Practice and Policy, 4(1), 74–83.

11. Bonanno, G. A., Mancini, A. D., Horton, J. L., Powell, T. M., Leardmann, C. A., Boyko, E. J., Wells, T. S., Hooper, T. I., Gackstetter, G. D., Smith, T. C., & Millennium Cohort Study Team. (2012). Trajectories of trauma symptoms and resilience in deployed U.S. military service members: prospective cohort study. The British Journal of Psychiatry: The Journal of Mental Science, 200(4), 317–323.

12. Bridgland, V. M. E., Moeck, E. K., Green, D. M., Swain, T. L., Nayda, D. M., Matson, L. A., Hutchison, N. P., & Takarangi, M. K. T. (2021). Why the COVID-19 pandemic is a traumatic stressor. PloS One, 16(1), e0240146.

13. Brown, G. W., Bifulco, A., Harris, T., & Bridge, L. (1986). Life stress, chronic subclinical symptoms and vulnerability to clinical depression. Journal of Affective Disorders, 11(1), 1–19.

14. Brülhart, M., Klotzbücher, V., Lalive, R., & Reich, S. K. (2021). Mental health concerns during the COVID-19 pandemic as revealed by helpline calls. Nature, 600(7887), 121–126.

15. Brunoni, A. R., Suen, P. J. C., Bacchi, P. S., Razza, L. B., Klein, I., Dos Santos, L. A., de Souza Santos, I., Valiengo, L. da C. L., Gallucci-Neto, J., Moreno, M. L., & Others. (2021). Prevalence and risk factors of psychiatric symptoms and diagnoses before and during the COVID-19 pandemic: findings from the ELSA-Brasil COVID-19 mental health cohort. Psychological Medicine, 1–12.

16. Caspi, A., & Moffitt, T. E. (2018). All for One and One for All: Mental Disorders in One Dimension. The American Journal of Psychiatry, 175(9), 831–844.

17. Chiba, T., Ide, K., Murakami, M., Kobayashi, N., Oka, T., Nakai, F., Yorizawa, R., Miyake, Y., Hamamura, T., Honjo, M., Toda, H., Kanazawa, T., Boku, S., Kubo, T., Hishimoto, A., Kawato, M., & Cortese, A. (2023). Event-related PTSD symptoms as a high-risk factor for suicide: longitudinal observational study. Nature Mental Health, 1(12), 1013–1022.

18. Cohen, J. A., & Mannarino, A. P. (2015). Trauma-focused cognitive behavior therapy for traumatized children and families. Child and Adolescent Psychiatric Clinics of North America, 24(3), 557–570.

19. deRoon-Cassini, T. A., Mancini, A. D., Rusch, M. D., & Bonanno, G. A. (2010). Psychopathology and resilience following traumatic injury: a latent growth mixture model analysis. Rehabilitation Psychology, 55(1), 1–11.

20. Dickstein, B. D., Suvak, M., Litz, B. T., & Adler, A. B. (2010). Heterogeneity in the course of posttraumatic stress disorder: trajectories of symptomatology: Trajectories of PTSD Symptomatology. Journal of Traumatic Stress, 23(3), 331–339.

21. Dohrenwend, B. P., Turner, J. B., Turse, N. A., Adams, B. G., Koenen, K. C., & Marshall, R. (2006). The psychological risks of Vietnam for U.S. veterans: a revisit with new data and methods. *Science (New York*, N.Y.), 313(5789), 979–982.

22. Foa, E. B., Kozak, M. J., Salkovskis, P. M., Coles, M. E., & Amir, N. (1998). The validation of a new obsessive–compulsive disorder scale: The Obsessive–Compulsive Inventory. Psychological Assessment, 10(3), 206–214.

23. Galatzer-Levy, I. R., Huang, S. H., & Bonanno, G. A. (2018). Trajectories of resilience and dysfunction following potential trauma: A review and statistical evaluation. Clinical Psychology Review, 63, 41–55.

24. Goh, C., & Agius, M. (2010). The stress-vulnerability model how does stress impact on mental illness at the level of the brain and what are the consequences? Psychiatria Danubina, 22(2), 198–202.

25. Harding, E. J., Paul, E. S., & Mendl, M. (2004). Cognitive bias and affective state. Nature, 427(6972), 312.

26. Harris, T. B., Carlisle, L. L., Sargent, J., & Primm, A. B. (2010). Trauma and diverse child populations. Child and Adolescent Psychiatric Clinics of North America, 19(4), 869–887.

27. Husky, M. M., Pietrzak, R. H., Marx, B. P., & Mazure, C. M. (2021). Research on posttraumatic stress disorder in the context of the COVID-19 pandemic: A review of methods and implications in general population samples. *Chronic Stress (Thousand Oaks*, Calif.), 5, 24705470211051327.

28. Judkins, J. L., Moore, B. A., Collette, T. L., Hale, W. J., Peterson, A. L., & Morissette, S. B. (2020). Incidence rates of posttraumatic stress disorder over a 17-year period in active duty military service members: PTSD incidence rates in service members. Journal of Traumatic Stress, 33(6), 994–1006.

29. Karstoft, K.-I., Armour, C., Elklit, A., & Solomon, Z. (2015). The role of locus of control and coping style in predicting longitudinal PTSD-trajectories after combat exposure. Journal of Anxiety Disorders, 32, 89–94.

30. Kessler, R. C., Adler, L., Ames, M., Demler, O., Faraone, S., Hiripi, E., Howes, M. J., Jin, R., Secnik, K., Spencer, T., Ustun, T. B., & Walters, E. E. (2005). The World Health Organization Adult ADHD Self-Report Scale (ASRS): a short screening scale for use in the general population. Psychological Medicine, 35(2), 245–256.

31. Kremer, L., Klein Holkenborg, S. E. J., Reimert, I., Bolhuis, J. E., & Webb, L. E. (2020). The nuts and bolts of animal emotion. Neuroscience and Biobehavioral Reviews, 113, 273–286.

32. Lantos, T., & Nyári, T. (2024). The impact of the first year of COVID-19 pandemic on suicides in a collection of 27 EU-related countries. Scientific Reports, 14. 10.1038/s41598-024-68604-3

33. Marx, B. P., Hall-Clark, B., Friedman, M. J., Holtzheimer, P., & Schnurr, P. P. (2024). The PTSD Criterion A debate: A brief history, current status, and recommendations for moving forward. Journal of Traumatic Stress, 37(1), 5–15.

34. Meerkerk, G.-J., Van Den Eijnden, R. J. J. M., Vermulst, A. A., & Garretsen, H. F. L. (2009). The Compulsive Internet Use Scale (CIUS): some psychometric properties. *Cyberpsychology & Behavior: The Impact of the Internet*, Multimedia and Virtual Reality on Behavior and Society, 12(1), 1–6.

35. Murphy, D., & Smith, K. V. (2018). Treatment efficacy for veterans with posttraumatic stress disorder: Latent class trajectories of treatment response and their predictors: Treatment trajectories for veterans with ptsd. Journal of Traumatic Stress, 31(5), 753–763.

36. Muthén, B. (2004). *Latent variable analysis. The Sage handbook of quantitative methodology for the social sciences*.

37. Muthén, B., & Muthén, L. K. (2000). Integrating person-centered and variable-centered analyses: growth mixture modeling with latent trajectory classes. Alcoholism, Clinical and Experimental Research, 24(6), 882–891.

38. Muysewinkel, E., Stene, L. E., Van Deynse, H., Vesentini, L., Bilsen, J., & Van Overmeire, R. (2024). Post-what stress? A review of methods of research on posttraumatic stress during COVID-19. Journal of Anxiety Disorders, 102(102829), 102829.

39. Nagamine, M., Giltay, E. J., Shigemura, J., van der Wee, N. J., Yamamoto, T., Takahashi, Y., Saito, T., Tanichi, M., Koga, M., Toda, H., Shimizu, K., Yoshino, A., & Vermetten, E. (2020). Assessment of factors associated with long-term posttraumatic stress symptoms among 56 388 first responders after the 2011 great east japan earthquake. JAMA Network Open, 3(9), e2018339.

40. Noworyta-Sokolowska, K., Kozub, A., Jablonska, J., Rodriguez Parkitna, J., Drozd, R., & Rygula, R. (2019). Sensitivity to negative and positive feedback as a stable and enduring behavioural trait in rats. Psychopharmacology, 236(8), 2389–2403.

41. Nylund-Gibson, K., & Choi, A. Y. (2018). Ten frequently asked questions about latent class analysis. Translational Issues in Psychological Science, 4(4), 440–461.

42. Nylund, K. L., Asparouhov, T., & Muthén, B. O. (2007). Deciding on the number of classes in latent class analysis and growth mixture modeling: A Monte Carlo simulation study. Structural Equation Modeling: A Multidisciplinary Journal, 14(4), 535–569.

43. Oka, T., Hamamura, T., Miyake, Y., Kobayashi, N., Honjo, M., Kawato, M., Kubo, T., & Chiba, T. (2021). Prevalence and risk factors of internet gaming disorder and problematic internet use before and during the COVID-19 pandemic: A large online survey of Japanese adults. Journal of Psychiatric Research, 142, 218–225.

44. Oka, T., Kubo, T., Kobayashi, N., Nakai, F., Miyake, Y., Hamamura, T., Honjo, M., Toda, H., Boku, S., Kanazawa, T., Nagamine, M., Cortese, A., Takebayashi, M., Kawato, M., & Chiba, T. (2021). Multiple time measurements of multidimensional psychiatric states from immediately before the COVID-19 pandemic to one year later: a longitudinal online survey of the Japanese population. Translational Psychiatry, 11(1), 573.

45. Orcutt, H. K., Erickson, D. J., & Wolfe, J. (2004). The course of PTSD symptoms among Gulf War veterans: a growth mixture modeling approach. Journal of Traumatic Stress, 17(3), 195–202.

46. Pan, L., Xu, Q., Kuang, X., Zhang, X., Fang, F., Gui, L., Li, M., Tefsen, B., Zha, L., & Liu, H. (2021). Prevalence and factors associated with post-traumatic stress disorder in healthcare workers exposed to COVID-19 in Wuhan, China: a cross-sectional survey. BMC Psychiatry, 21(1), 572.

47. Pierce, M., McManus, S., Hope, H., Hotopf, M., Ford, T., Hatch, S. L., John, A., Kontopantelis, E., Webb, R. T., Wessely, S., & Abel, K. M. (2021). Mental health responses to the COVID-19 pandemic: a latent class trajectory analysis using longitudinal UK data. The Lancet. Psychiatry, 8(7), 610–619.

48. Pirkis, J., John, A., Shin, S., DelPozo-Banos, M., Arya, V., Analuisa-Aguilar, P., Appleby, L., Arensman, E., Bantjes, J., Baran, A., Bertolote, J. M., Borges, G., Brečić, P., Caine, E., Castelpietra, G., Chang, S.-S., Colchester, D., Crompton, D., Curkovic, M., … Spittal, M. J. (2021). Suicide trends in the early months of the COVID-19 pandemic: an interrupted time-series analysis of preliminary data from 21 countries. The Lancet. Psychiatry, 8(7), 579–588.

49. Radloff, L. S. (1977). The CES-D Scale: A Self-Report Depression Scale for Research in the General Population. Applied Psychological Measurement, 1(3), 385–401.

50. Saito, T., van der Does, F. H. S., Nagamine, M., van der Wee, N. J., Shigemura, J., Yamamoto, T., Takahashi, Y., Koga, M., Toda, H., Yoshino, A., Vermetten, E., & Giltay, E. J. (2022). Risk and resilience in trajectories of post-traumatic stress symptoms among first responders after the 2011 Great East Japan Earthquake: 7-year prospective cohort study. The British Journal of Psychiatry: The Journal of Mental Science, 221(5), 1–8.

51. Saunders, J. B., Aasland, O. G., Babor, T. F., de La Fuente, J. R., & Grant, M. (1993). Development of the Alcohol Use Disorders Identification Test (AUDIT): WHO Collaborative Project on Early Detection of Persons with Harmful Alcohol Consumption-II. Addiction, 88(6), 791–804.

52. Schäfer, S. K., Kunzler, A. M., Kalisch, R., Tüscher, O., & Lieb, K. (2022). Trajectories of resilience and mental distress to global major disruptions. Trends in Cognitive Sciences, 26(12), 1171– 1189.

53. Solomon, Z., Mikulincer, M., Ohry, A., & Ginzburg, K. (2021). Prior trauma, PTSD long-term trajectories, and risk for PTSD during the COVID-19 pandemic: A 29-year longitudinal study. Journal of Psychiatric Research, 141, 140–145.

54. Spielberger, C. D. (1983). State-Trait Anxiety Inventory for Adults. 10.1037/t06496-000

55. Steenkamp, M. M., Dickstein, B. D., Salters-Pedneault, K., Hofmann, S. G., & Litz, B. T. (2012). Trajectories of PTSD symptoms following sexual assault: is resilience the modal outcome?: Trajectories of PTSD Following Sexual Assault. Journal of Traumatic Stress, 25(4), 469–474.

56. van der Velden, P. G., Hyland, P., Contino, C., von Gaudecker, H.-M., Muffels, R., & Das, M. (2021). Anxiety and depression symptoms, the recovery from symptoms, and loneliness before and after the COVID-19 outbreak among the general population: Findings from a Dutch population-based longitudinal study. PloS One, 16(1), e0245057.

57. Vapnik, V., Golowich, S., & Smola, A. (1996). Support Vector method for function approximation, regression estimation and signal processing. Neural Information Processing Systems, 281–287.

58. Weiss, D. S. (2004). The Impact of Event Scale: Revised. In Cross-Cultural Assessment of Psychological Trauma and PTSD (pp. 219–238). 10.1007/978-0-387-70990-1_10

59. Wold, S., Esbensen, K., & Geladi, P. (1987). Principal component analysis. Chemometrics and Intelligent Laboratory Systems, 2(1), 37–52.

