## Supplemental Figures and Tables for "Pre-event psychiatric states predict trajectories of stress symptoms in Japan"

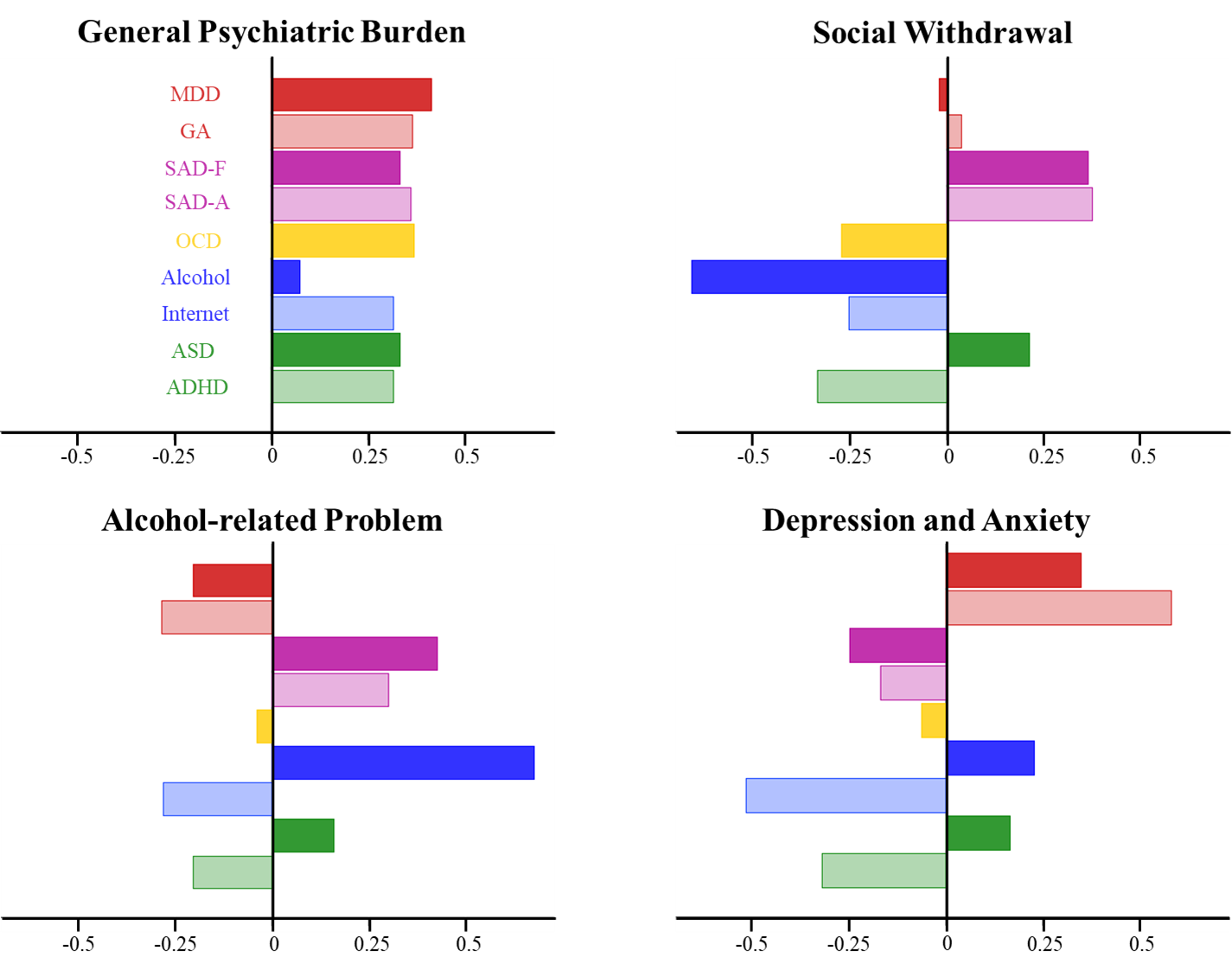

**Figure S1. Loadings on each principal component.**

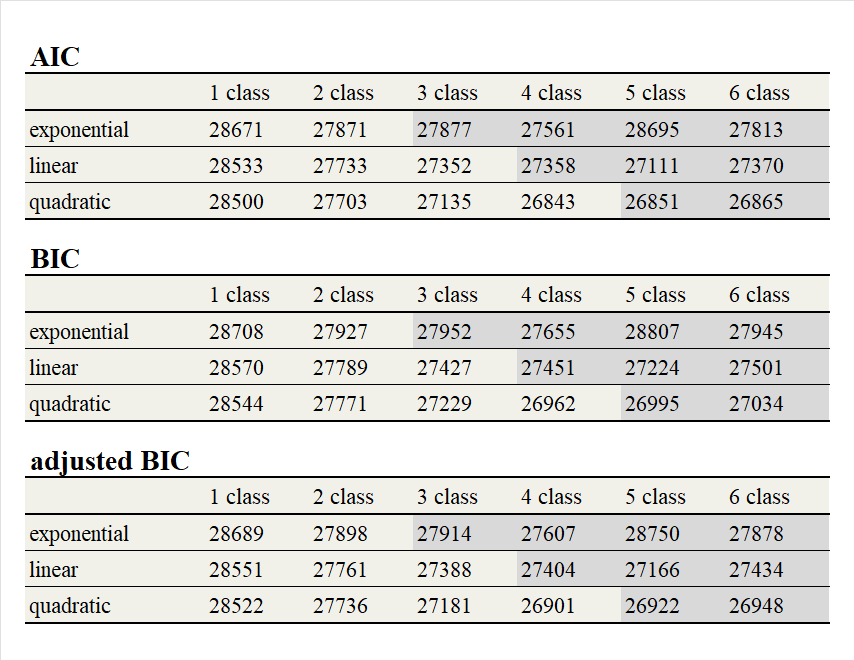

**Table S1. Evaluation of the LGMM fitting.**

All three indicators showed [4 class] - [quadratic] condition was the optimal classification parameter in the latent growth mixture analysis. The shaded cell condition indicates bad classification (including classes that had no individuals assigned or no converged).

**Statistical analysis of principle components (PCs) trajectories**

In order to test the interaction between modifying factors and symptoms, the temporal change of PC for each trajectory group was tested by unpaired t-test (multiple comparisons were corrected using the Bonferroni method). As a result, a significant increase of the general psychiatric burden component at T1 from T0 was observed in the mild chronic (*p* <.001, *t* = 4.70, df = 1190), early response (*p* <.001, *t* = 5.18, df = 710) and resilient groups (*p* <.001, *t* = 4.81, df = 5188). A significant decrease in the general psychiatric burden component was also observed at T2 from T1 in the early response group (*p* <.01, *t* = 4.03, df = 579) and at T5 from T4 in the resilient group (*p* <.001, *t* = 3.99, df = 2805). A significant change in the social withdrawal component was only observed in the resilient group at T1 from T0 (increase, *p* <.001, *t* = 6.09, df = 5188) and T5 from T4 (decrease, *p* <.001, *t* = 4.62, df = 2805). A significant change in the depression/anxiety component was observed in the resilient group at T1 from T0 (increase, *p* <.001, *t* = 4.64, df = 5188). Temporal change from T0 was also tested by unpaired *t*-test (multiple comparison was corrected using the Bonferroni method, see Table S2).

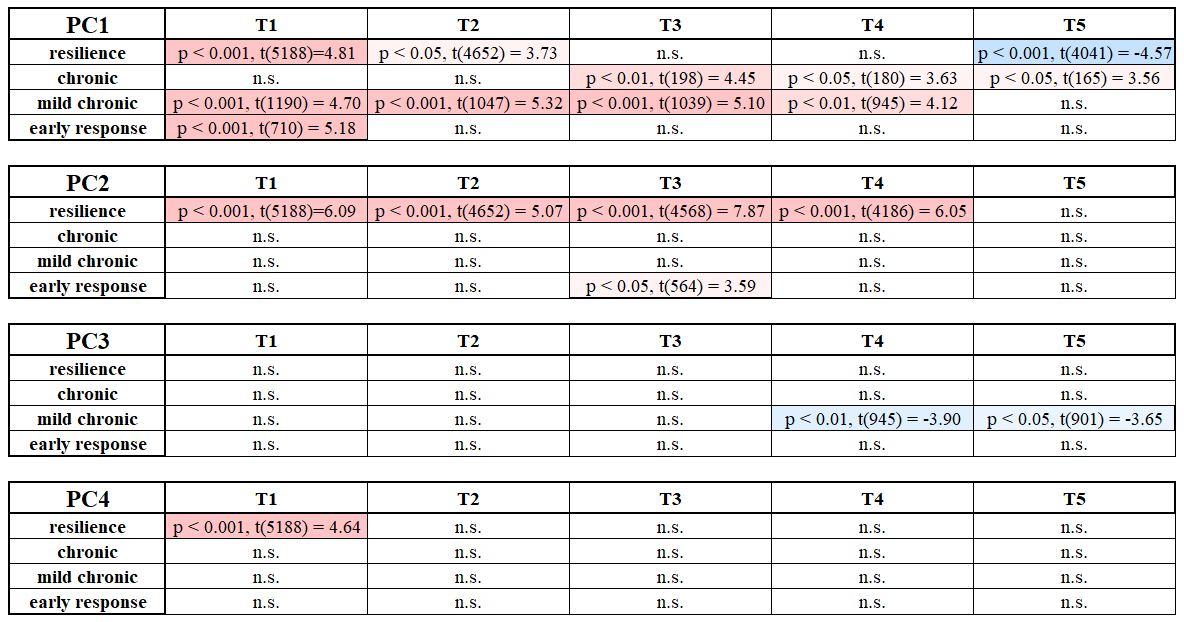

**Table S2.** Significant difference between latent stress symptoms and each time point for each PC.

**Leave-one-time-point-out cross-validation analysis**

The leave-one-time-point-out cross-validation analysis showed higher than 0.5 adjusted-R^2^ score for all Tx conditions at training and higher than 0.4 adjusted-R^2^ score for all Tx conditions at test (T1: training; adjusted-R^2^ = 0.505, testing; adjusted-R^2^ = 0.451, T2: training; adjusted-R^2^ = 0.543, testing; adjusted-R^2^ = 0.374, T3: training; adjusted-R^2^ = 0.543, testing; adjusted-R^2^ = 0.435, T4: training; adjusted-R^2^ = 0.530, testing; adjusted-R^2^ = 0.414, T5: training; adjusted-R^2^ = 0.529, testing; adjusted-R^2^ = 0.405, averaged: training; adjusted-R^2^ = 0.530, testing; adjusted-R^2^ = 0.416).

**Stepwise regression analysis for reliability estimation of the latent stress symptoms**

In order to apply the latent stress symptoms estimation easily in a clinical setting, stepwise regression was performed to clarify the number of psychological disorder questionnaires required to reliably estimate the stress symptom score. This analysis consisted of two steps. The first was the ordering step and the second was the regression step. In the ordering step, data were analyzed by multiple regression. Questionnaires were ordered according to coefficients and the lowest item was rejected. This process was repeated until all questionnaires had been rejected and then, questionnaires were ordered according to the order in which they remained un-rejected. The ordering step was repeated 100 times and the averaged order of questionnaires was estimated. In the regression step, half of the data was assigned to the training dataset and the other half to the test dataset. The regression model was trained by multiple regression, and the regression was performed on the top 3 (CESD, OCI and CIUS) or 6 (CESD, OCI, CIUS, AQ, STAI and AUDIT) questionnaires, according to the averaged order. The whole procedure from dataset separation to parameter estimation was repeated 100 times and accuracy was evaluated using the averaged adjusted-R^2^ (6 var.: training; adjusted-R^2^ = 0.439, test; adjusted-R^2^ = 0.437, 3 var.: training; adjusted-R^2^ = 0.433, test; adjusted-R^2^ = 0.432).
